# Perceptions of Optimal Post-Malnutrition Weight Gain & Growth: a global mixed methods study

**DOI:** 10.1101/2025.07.11.25331352

**Authors:** Grace O’Donovan, Kelda Septima Yeung, Mubarek Abera, Elizabeth Wimborne, Gemechu Ameya, Amir Kirolos, Laurentya Olga, Kimberley McKenzie, Albert Koulman, Debbie S. Thompson, Jonathan Swann, CHANGE project, Marko Kerac

## Abstract

**Introduction:** The treatment of severe childhood malnutrition focuses on short-term outcomes, like mortality prevention and anthropometric recovery. However, emerging evidence suggests a possible association between post-malnutrition growth and later non-communicable disease (NCD) risk. Our aim was to inform future child malnutrition treatment programmes by describing perceptions of optimal rates of post-malnutrition weight gain/growth; assessing how short- and long-term outcomes are currently understood and prioritised; understanding perceptions of the role of malnutrition treatment services in preventing longer-term NCDs.

**Methods:** A mixed methods study, involving a global cross-sectional online survey (December 2023-March 2024) and key informant interviews (March-July 2024). Participants were professionals with experience in severe malnutrition and/or child health, identified through convenience and snowballing sampling. Quantitative data were analysed by descriptive statistics, while qualitative data were explored by thematic analysis.

**Results:** Sixty-eight survey respondents were included, from a range of backgrounds. Ten were also interviewed. Almost half of survey participants perceived 5-10 g/kg/d as optimal weight gain in both inpatient and outpatient treatment. In terms of programme aims, 71% ranked ‘preventing mortality’ as the most important. Two-thirds (66%) rated reducing the risk of adulthood NCDs as a ‘very important’ long-term aim, while 3% said this was of ‘low/no importance’. Slower post-malnutrition weight gain was seen as beneficial by 69% of respondents, while 13% had opposing views and 18% were unsure. Key concerns were avoiding overfeeding/physiological disturbances and radical changes among those who supported slower weight gain, and worries about not meeting energy requirements among those who did not.

**Conclusions:** There is strong consensus that preventing mortality remains the key aim of malnutrition treatment programmes but also increasing recognition of a potential to impact long-term NCD risk. Towards this, perceptions of optimal post-malnutrition weight gain targets vary, and better evidence is urgently needed to inform future policy/practice.

**What is already known on this topic:** Treatment of severe childhood malnutrition focuses on short-term outcomes, such as mortality prevention and anthropometric recovery. Longer-term outcomes are rarely considered, yet recent evidence suggests a possible association between rapid post-malnutrition growth and later non-communicable disease (NCD) risk.

**What this study adds:** Policy makers, programme managers, researchers, and other professionals leading current nutrition treatment programmes prioritise short-term outcomes of malnutrition treatment but most also believe that treatment can impact long-term outcomes, including NCDs. However, perceptions of the importance of any impact vary, as do ideas about how this can be achieved and concerns regarding risk-benefit balance (*e.g.,* slower growth might reduce future NCD risk but compromise short-term outcomes like mortality).

**How this study might affect research, practice or policy:** Varied opinions on post-malnutrition weight gain and growth reflect large current evidence gaps and the urgent need for future research to determine optimal weight gain targets and growth patterns. Our data also highlights the need for greater consideration of long-term outcomes when planning malnutrition-related programmes and policy.

## INTRODUCTION

The double burden of malnutrition is the coexistence of undernutrition alongside overnutrition and diet-related non-communicable diseases (NCDs) [1]. Sustainable Development Goals (SDGs) highlight undernutrition and NCDs separately in SDG 2 and SDG 3 Target 4 [2]. However, there is growing evidence to suggest that undernutrition in early life, especially when followed by overweight/obesity, predisposes to adulthood NCDs [3]. Recognising this, the World Health Organisation (WHO) calls for new integrated approaches involving double-duty actions that simultaneously tackle both extremes of malnutrition [1].

Childhood undernutrition confers high risks of mortality, contributing to the deaths of some three million children aged under five each year [4]. Malnutrition treatment programmes often strive to maximise rates of post-malnutrition weight gain (PMWG) and growth with the immediate interests of preventing mortality and achieving anthropometric targets. However, emphasis on short-term outcomes can overlook potential long-term impacts. Severe childhood malnutrition affects survivors into adulthood with studies indicating an association between childhood undernutrition and increased risks of cardiometabolic NCDs [5, 6]. These findings are consistent with the “Developmental Origins of Health and Disease” (DOHaD) hypothesis [7] and the capacity-load model, in which childhood undernutrition reduces metabolic capacity, thus increasing vulnerability to later NCD [8]. Among those exposed to unhealthy obesogenic environments in later life, NCD risk may be particularly exacerbated among those who previously experienced child undernutrition. More importantly, the evidence from studies of childhood malnutrition indicates that the period during which developmental plasticity shapes long-term capacity for homeostasis extends beyond the pre-natal period [5]. Data from high-income countries suggest that excessive PMWG might play a mechanistic role contributing to adulthood NCDs [9]. Very rapid PMWG in childhood has been associated with higher adiposity in adult survivors in Jamaica [10] and slower patterns of weight gain seem more conducive with lower cardiometabolic risks in later life [11].

High prevalence of undernutrition in low- and middle-income settings may contribute to NCD burden [12] and all possible actions to lower the risks are important. Better managing PMWG may be one approach [13]. We hypothesise that current malnutrition treatment programmes have the potential to optimise rates of PMWG, achieving desirable short-term outcomes whilst simultaneously minimising long-term morbidity for survivors of severe malnutrition. Towards this, it is important to understand how health professionals interpret different patterns of PMWG as plotted on clinical charts which are widely used to manage growth [14, 15].

Our aim is to inform future research, policy, and practice by understanding current perceptions of childhood PMWG and its relation to adult NCD risk. Specific objectives are:

- to describe key stakeholders’ perceptions of optimal rates of PMWG and catch-up growth;
- to assess how short- and long-term outcomes are understood and prioritised in current malnutrition treatment programmes;
- to identify stakeholders’ perceptions of the role of malnutrition treatment services in preventing long-term risk of adulthood NCDs.

## METHODS

This mixed-methods study explored perceptions of childhood PMWG and growth, and their relation to long-term NCD risk. The study primarily consisted of an online cross-sectional survey, complemented by qualitative in-depth key informant interviews. We reported according to the Checklist for Reporting of Survey Studies (CROSS) (Appendix A) [16].

### Survey

The survey was open on Jisc Online Surveys v3 between 13/12/2023 and 19/03/2024 [17]. Participants were asked about optimal weight gain during inpatient and outpatient care. Then, they compared hypothetical growth charts, designed to represent fast vs slower growth, with both weight-for-age z-score (WAZ) and height-for-age z-score (HAZ) comparisons. Lastly, participants were asked about short- and long-term aims of malnutrition treatment programmes. The questionnaire (Supplementary materials) was designed to be completed in approximately 15 minutes. Healthcare professionals with experience in severe malnutrition and/or child health were eligible to participate, enabling a wide range of responses from different settings.

Participants were recruited through professional networks and existing online platforms (Health Information for All and en-net.org). Additional snowball sampling involved distributing the invitation through our professional networks and encouraging participants to share the invitation through their own professional networks. There was no prespecified sample size, as this is an exploratory study. We examined the list of consenting participants to ensure there were no duplicate entries or events of multiple participation. We analysed responses using descriptive statistics on Excel and STATA SE/18. We then performed thematic analyses of short, open-ended answers to identify common themes.

### Interviews

Survey participants were given the option to provide further consent and their contact details if they were willing to be invited for in-depth interview. This online interview was designed to last 30-45 minutes. This semi-structured, key informant interview followed a topic guide developed by lead investigators. It included open-ended, non-leading questions to encourage elaboration from participants. As the purpose of the interviews was to complement survey findings, there was no pre-defined sample size. However, we projected to recruit at least 10 participants, as an estimate for approaching data saturation [18, 19]. Participants were purposively selected, with the intention of capturing a variety of opinions from different backgrounds.

Participants identified for interview were emailed with an invitation, information sheet, and consent form. Once consent had been obtained, G.O.D. (MSc), a female Research Assistant, conducted the interviews. Only the participant and G.O.D. were present during the interview (apart from one interview which M.K. attended). Participants were given the option of audio-only or audio-visual Zoom call. With consent, the interviews were recorded and transcribed. Participants had the option of reviewing their interview transcript upon request. Interviews were conducted between March and July 2024. We analysed interview transcripts for emerging themes and identified quotations that complemented our findings.

### Ethical considerations

This study received ethics approval from the London School of Hygiene and Tropical Medicine (LSHTM) Research Ethics Committee (Reference: 26610). Informed consent was collected separately for both survey and interview participation. Only survey respondents willing to be contacted for interview were asked to provide their email address, for the purposes of obtaining informed consent and scheduling the interview. Once interview-specific written consent had been received and the interview had been scheduled, all contact details and email correspondence were deleted from our records to maintain privacy and confidentiality. This low-risk project collected non-sensitive and non-personal data. Through the Jisc Online Survey platform, survey data was processed according to strict information security standards. Interview recordings were erased once transcription was complete. Survey data and interview transcripts were stored securely in a password-protected, encrypted folder for the duration of the study and were added to the LSHTM Data Compass Repository. All published data and results were anonymised and de-identified.

## RESULTS

### Participants’ characteristics

Of 79 survey respondents, 68 gave consent for inclusion (Figure 1). Of these, 31 gave consent for invitation to interview. Twenty-one were invited for interview. Ten consented to and attended interview. Table 1 shows characteristics of survey respondents. Almost half were nutritionists (46%). Most were working in Non-Governmental Organisations/charity (37%) and/or academia (35%). Most respondents’ work (62%) focussed on Africa, 16% on Latin America and the Caribbean, 13% on Asia, and 12% on Europe.

**Figure 1.**
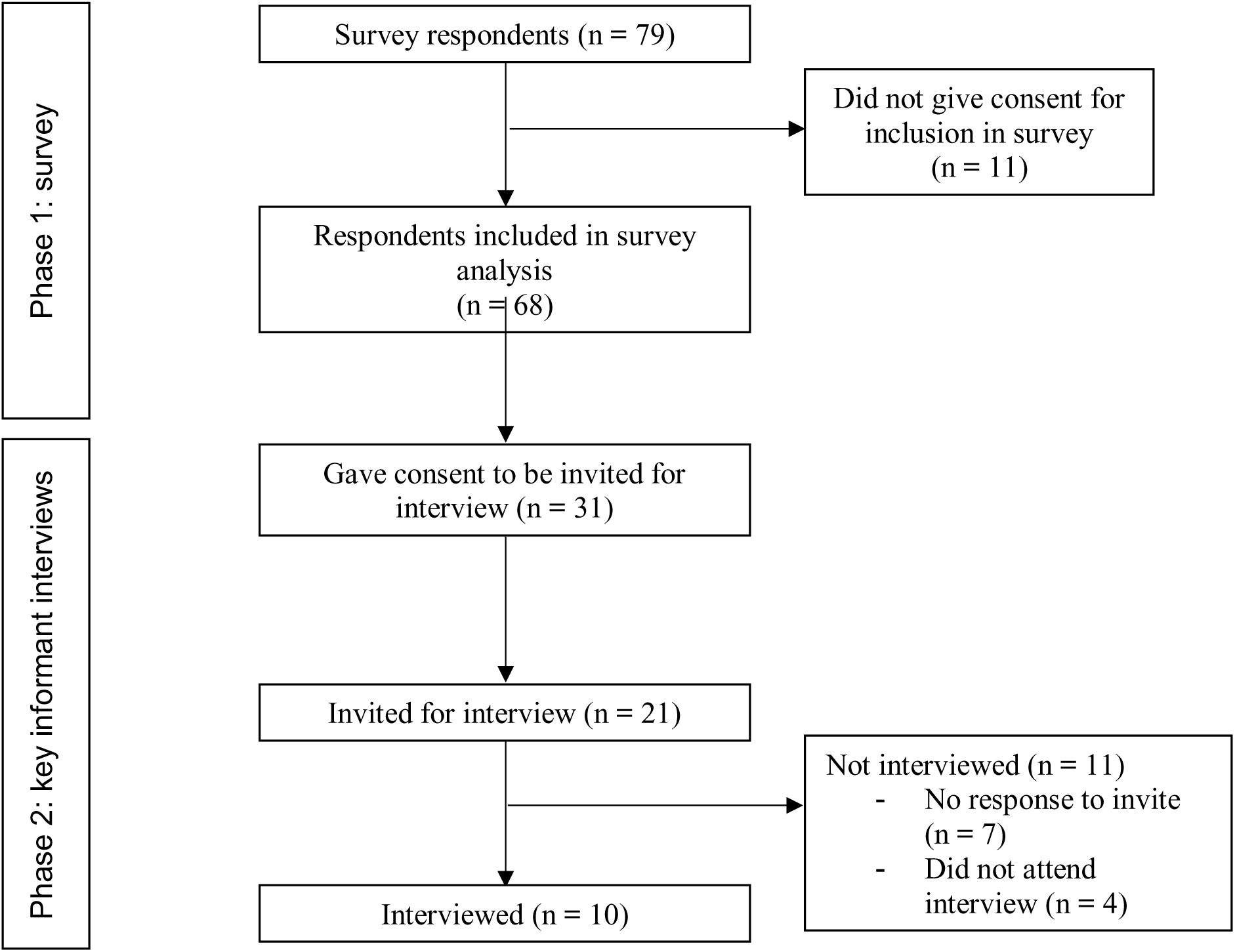
Participant flow diagram (N=79)

**Table 1.** Characteristics of survey respondents included in this study (N=68)

|  |  | Number of participants | Percentage (%) |
| --- | --- | --- | --- |
| Organisation | Non-Governmental Organisation/Charity | 25 | 37 |
|  | Academic | 24 | 35 |
|  | Healthcare | 7 | 10 |
|  | Government | 4 | 6 |
|  | United Nations | 4 | 6 |
|  | Public Health | 2 | 3 |
|  | Other* | 2 | 3 |
| Role | Nutritionist | 31 | 46 |
|  | Public Health Specialist | 14 | 21 |
|  | Nurse | 9 | 13 |
|  | Programme Manager | 7 | 10 |
|  | Doctor (paediatrics) | 6 | 9 |
|  | Doctor (other) | 5 | 7 |
|  | Other<sup>†</sup> | 18 | 27 |
| Context of work | Humanitarian | 37 | 54 |
|  | Development | 29 | 43 |
|  | Other<sup>‡</sup> | 21 | 31 |
| Focus region of work | Sub-Saharan Africa | 41 | 60 |
|  | Global | 12 | 18 |
|  | Latin America & Caribbean | 11 | 16 |
|  | Europe | 8 | 12 |
|  | South-East Asia | 5 | 7 |
|  | South Asia | 5 | 7 |
|  | East Asia | 3 | 4 |
|  | North America | 3 | 4 |
|  | West Asia | 2 | 3 |
|  | North Africa | 1 | 2 |
|  | Oceania | 1 | 2 |
| Role length | 0-5 years | 20 | 29 |
|  | 6-10 years | 16 | 24 |
|  | 11-15 years | 10 | 15 |
|  | 16-20 years | 4 | 6 |
|  | 20+ years | 18 | 27 |

\*Both participants who selected “Other” for their organisation indicated they are students.
<sup>†</sup> A range of roles were indicated by those who selected “Other” for their role: researcher/scientist, dietitian, student, programme coordinator, public health officer, healthcare worker, executive director, health and nutrition supervisor.
<sup>‡</sup> A range of contexts were indicated by those who selected “Other”: mainly research and education, but also health/public health/healthcare, clinical medicine, programme implementation in schools, infectious and tropical diseases, food security, and emergency response.

### Perceptions of optimal rates of PMWG

Table 2 indicates participants’ perceptions of PMWG. Figure 2 shows the responses when participants were asked their preference between two growth charts at a time. Most participants gave explanations for their preferences, ranging from 84% (57/68) for the Chart A vs B comparison to 72% (49/68) for Chart J vs K. Chart A vs B (designed as a ‘warm-up’, least controversial scenario) was the least divisive comparison, with 87% (59/68) preferring Chart A. Two main themes explained this preference: growth in Chart B tapered/plateaued towards the end (35/50), and Chart A increased with the median (20/50).

**Figure 2.**
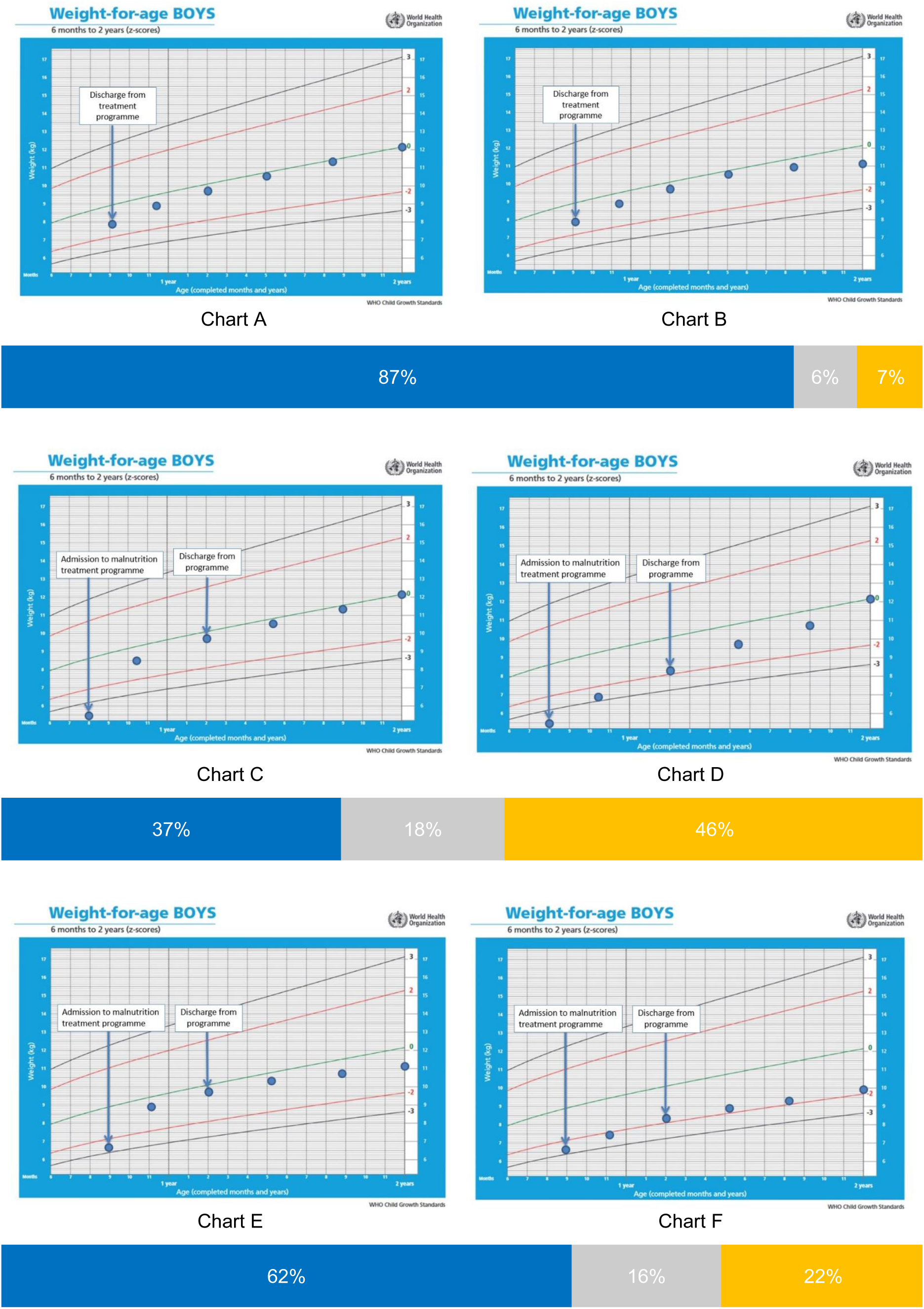
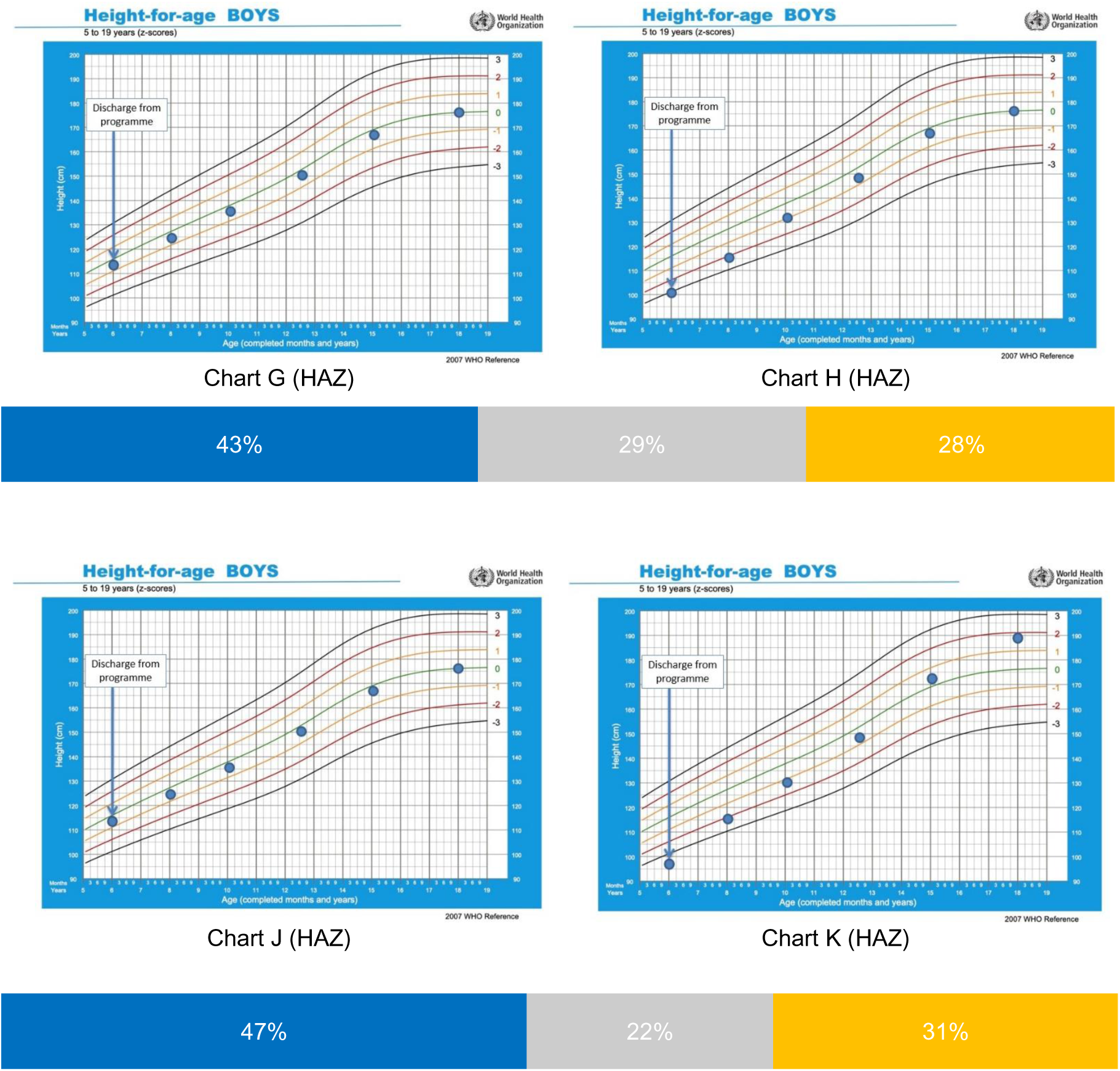
Participants’ responses when asked to indicate which growth chart showed a more optimal (healthier/more desirable) growth pattern, comparing two growth charts at a time (N=68) Note: Blue bar indicates preference for left-hand chart, orange indicates preference for right-hand chart, grey indicates “no difference”. Charts A-F are weight-for-age z-scores and Charts G-K are height-for-age z-scores.

**Table 2.** Participants’ perceptions of post-malnutrition weight gain and aims of child malnutrition treatment programmes (N=68)

|  |  | Number of participants | Percentage (%) |
| --- | --- | --- | --- |
| Optimal weight gain during inpatient phase of treatment (g/kg/d) | 0-5 | 15 | 22 |
|  | 5-10 | 33 | 49 |
|  | 10-15 | 17 | 25 |
|  | 15-20 | 2 | 3 |
|  | 20+ | 1 | 2 |
| Optimal weight gain during outpatient phase of treatment/CMAM (g/kg/d) | 0-5 | 14 | 21 |
|  | 5-10 | 32 | 47 |
|  | 10-15 | 16 | 24 |
|  | 15-20 | 5 | 7 |
|  | 20+ | 1 | 2 |
| Optimal weight gain during inpatient phase relative to outpatient phase/CMAM | Higher | 21 | 31 |
|  | Equivalent | 25 | 37 |
|  | Lower | 22 | 32 |
| Importance of "improving child development" as a medium-term aim of programmes | Very important | 54 | 79 |
|  | Important | 12 | 18 |
|  | Moderately important | 2 | 3 |
|  | Of low importance | 0 | 0 |
|  | Not at all important | 0 | 0 |
| Importance of "reducing the risk of NCDs in adulthood" as a long-term aim of programmes | Very important | 45 | 66 |
|  | Important | 17 | 25 |
|  | Moderately important | 4 | 6 |
|  | Of low importance | 1 | 2 |
|  | Not at all important | 1 | 2 |
| Belief that programmes have a role in reducing risk of adulthood NCDs, given that children survive childhood malnutrition | Yes | 60 | 88 |
|  | No | 3 | 4 |
|  | Don't know | 5 | 7 |
| Belief that slower rate of daily weight gain and growth post-malnutrition (*e.g.*, by providing lower-energy therapeutic feed) could be beneficial for the child in any way | Yes | 47 | 69 |
|  | No | 9 | 13 |
|  | Don't know | 12 | 18 |

NCDs=Non-Communicable Diseases; CMAM=Community Management of Acute Malnutrition

Chart C vs D (designed to represent faster vs slower weight recovery) was the most divisive WAZ growth chart comparison, with 46% (31/68) preferring Chart D, 37% (25/68) preferring Chart C, and 18% (12/68) selecting “no difference”. Nine explained their preference for Chart D due to dislike of rapid weight gain (9/22), with four specifying that this could have negative long-term effects. Six participants (6/22) indicated their belief that slow/steady/gradual growth was preferable. Two main themes emerged explaining preference for Chart C: faster recovery/rapid weight gain out of malnutrition (8/23) and aligning more closely with the median (8/23). Four participants (4/23) mentioned that Chart D continuing to cross centiles could result in overweight. The main themes given as reasons for selecting “no difference” were that both charts showed positive growth (5/10) and had the same end result (4/10).

Most (62%) preferred Chart E over F (designed to represent faster initial weight recovery vs slower weight recovery along a lower WAZ, respectively). The most common themes explaining this preference were that Child F remained malnourished/close to malnutrition (14/34), and Child E was closer to the median (10/34). The most common reason for preferring Chart F was consistent/steady growth (7/11). Four participants (4/11) suggested an indication of growth faltering in Chart E. Of nine that explained selecting “no difference”, three balanced growth faltering in Chart E with borderline WAZ in Chart F. Across those that chose Chart F and “no difference”, five participants (5/20) wanted to know the length/height.

In Chart G vs H (designed to represent post-discharge HAZ pattern starting from normal HAZ vs stunted, respectively, and both ending at HAZ=0), 43% (29/68) preferred Chart G, 28% (19/68) preferred Chart H, and 29% (20/68) selected “no difference”. The main themes among participants that explained preferring Chart G were that growth was more aligned with the median (10/22), and that the child was not discharged with an anthropometric deficit (unlike in Chart H) (10/22). Of 11 that explained their preference for Chart H, 10 participants alluded to the fact that there was a large improvement in Chart H. Of 18 that explained selecting “no difference”, eight mentioned that both charts achieved the same/normal HAZ.

Almost half (47%; 32/68) preferred Chart J (post-discharge HAZ consistently in the normal range), while 31% (21/68) preferred Chart K (designed to represent post-discharge HAZ regain from HAZ<-3 to HAZ>1), and 22% (15/68) found “no difference”. Of 25 that explained preferring Chart J, 13 explained this was because the growth was normal/consistent with the curve. Eleven suggested that growth in Chart K was very/too rapid. Four preferred Chart J because the child was never stunted. Four participants questioned the data validity in Chart K. Of 13 that explained choosing Chart K, nine reasoned that this showed faster growth. Of 11 who explained selecting “no difference”, four wanted more information. Three mentioned that both charts ended in the normal range.

### Understanding of short- and long-term outcomes

Regarding malnutrition treatment programmes, 66% thought that reducing adulthood NCD risk was a very important long-term aim (Table 2). Only two respondents (3%) thought this was of low/no importance. When ranking five aims in priority order (Figure 3), 71% ranked preventing mortality as the most important. However, 22% ranked improving child development as the most important. While some interview participants discussed the association between early life undernutrition and cognitive issues, there were mixed views on the potential impact of treatment programmes on cognitive outcomes, with some thinking this was asking too much, while others believed this to be an opportunity to improve neurodevelopment. Almost half (46%) ranked reducing the risk of short-term morbidity as the second most important aim.

**Figure 3.**
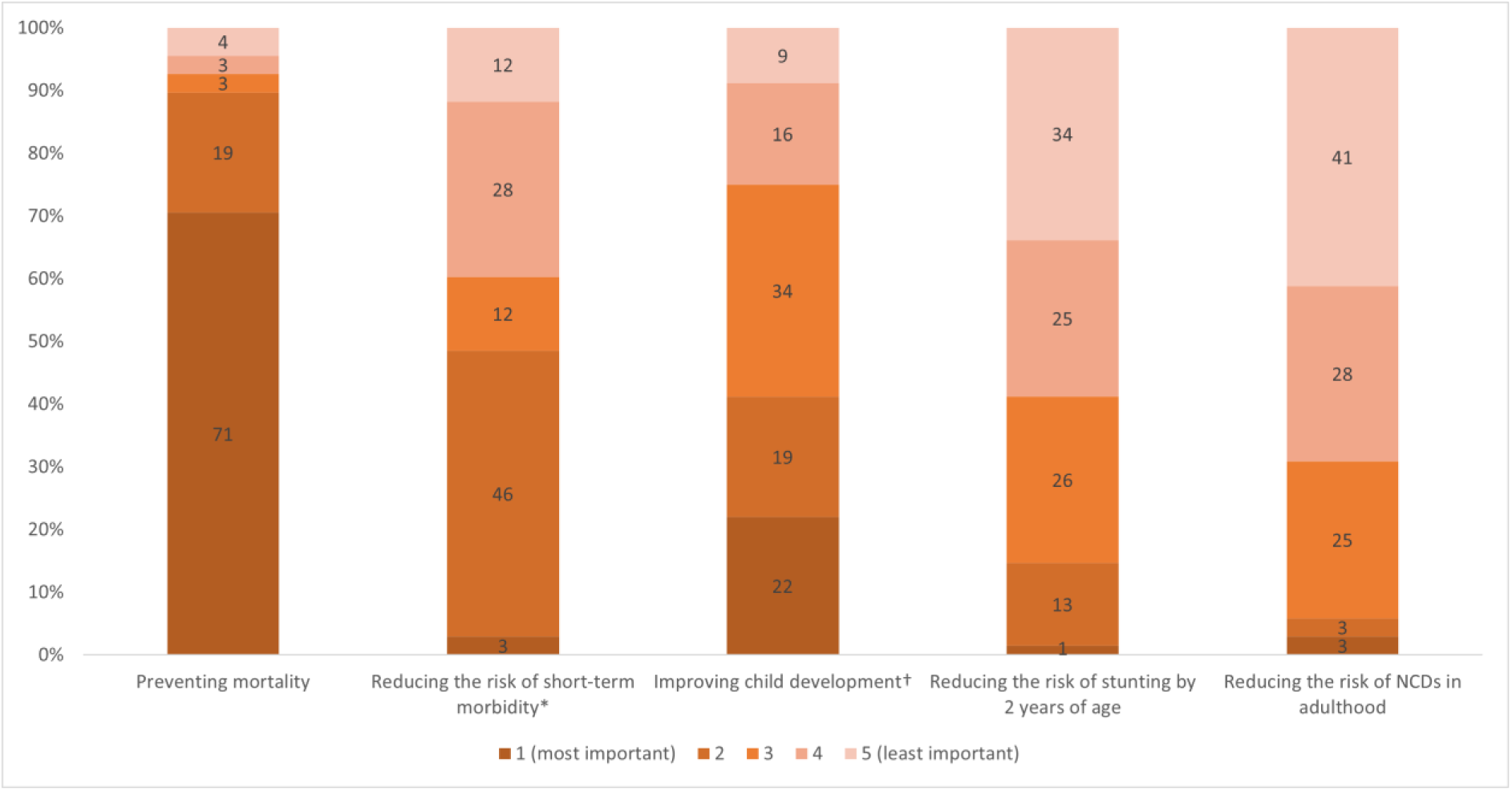
Participants’ responses (%) when asked to rank the importance and priority of five potential aims of malnutrition treatment programmes (N=68) *Short-term morbidity, *e.g.*, diarrhoea, pneumonia, acute infections, or illnesses ^†^Improving child development may include improving educational potential and preventing disability

### Perception of the role of childhood malnutrition treatment in preventing long-term NCD risk

Most respondents (88%) believed that programmes have a role in reducing later NCD risk in survivors, with 68% (41/60) providing an explanation. Fifteen participants (15/41) referred to the link between childhood undernutrition and later NCD risk. Nine (9/41) alluded to reductive adaptation as the link between childhood undernutrition and long-term outcomes (*e.g.*, the DOHaD hypothesis). *“Children who suffer and survive undergo many physiologic adaptations including vascular changes, hormonal changes and enzymatic changes during their development which may put them at risk of NCD later in their life.”* P58 survey response. Four (4/41) referred to links via socio-economic factors (*e.g.*, educational attainment, earning potential). Five participants (5/41) mentioned that rapid weight gain has been associated with later NCDs. “*There is growing evidence that both malnutrition and rapid weight gain during recovery may increase the risk of later-life NCDs.”* P03 survey response.

When asked if slower daily PMWG and growth (*e.g.*, by prescribing feeds at the lower end of the WHO-recommended range, *i.e.*, 150 kcal/kg/d) could be beneficial in any way, the majority answered “yes” (69%), but 18% were unsure, and 13% answered “no”. Of the 30 that explained why they answered “yes”, six participants thought it could be beneficial by avoiding overfeeding/physiological disturbances. Four participants (4/30) alluded to the idea that avoiding radical changes is better. Three (3/30) spoke about slower change being more sustainable. Three (3/30) mentioned avoiding excess fat accumulation.

Two participants expressed concerns over the current recommended dosage from WHO: *“I think there’s a problem with the estimation of energy requirement…so I think we give too much. So I think definitely we should reduce, especially at the end of the treatment.”* P01 interview response. A participant referred to evidence that reduced dosages *“are non-inferior to the standard ones in terms of recovery rate, mortality rate, and other factors, while also reaching more children”* P47 survey response. One survey participant mentioned that reduced dosage *“may not necessarily result in significantly lower weight gains”* P31 survey response. Another suggested that healthcare workers usually provide therapeutic feed at *“the higher quantity”* when given a range (P14 interview response). One participant spoke about the practicalities of treatment: *“this level of “technicality” may be superficial because it will be very challenging to implement”* P42 survey response.

Many participants thought slow growth would be preferable: “*the whole body and the organs are in the status of malfunction…they need time to recovery: not only the growth, not only the weight, but also the function.”* P20 interview response. Someone highlighted the need to understand the mechanism potentially linking weight recovery rate to NCD risk: *“When we do a slow weight gain, what body activities are we triggering so that in adulthood we don’t have these non-communicable diseases”* P14 interview response.

Of 12 participants that explained their selection of “don’t know”, two commented on limited data, two indicated preferring local diet, and one was concerned about long length of stay. Of four participants that explained why they disagreed, three talked about needing to meet energy requirements. There was concern over reducing therapeutic feed to get slower weight gain “*because of the potential association with mortality”* P01 interview response. There was scepticism over the link between weight gain and NCD risk: *“It’s not the rapid weight gain itself. It’s the underlying, other physiological problems that then link to NCDs.”* P03 interview response.

## DISCUSSION

There is a lack of consensus among key stakeholders on the optimal rate of PMWG. Most prioritise short-term outcomes (*i.e.*, reducing mortality and morbidity), with many focusing on rapid weight recovery to achieve this. However, many also recognised potential implications for long-term outcomes, with 88% agreeing that malnutrition treatment programmes have a role in reducing risk of adulthood NCDs in survivors and 69% believing that slower PMWG could be beneficial in some way, although opinions varied on whether slower PMWG could reduce later NCD risk. Some participants were concerned about mortality risk and that there was insufficient evidence for a causal link between rapid PMWG and NCD risk. On the other hand, some participants believed that slower weight gain would help the body to transition back to normal function, and that it would avoid continuing anthropometric increase towards overweight.

Almost half selected 5-10 g/kg/d as a desired optimal inpatient and outpatient weight gain, consistent with the WHO guidelines (2023) that used this as a target weight gain [20]. Only 21% believed 0-5g/kg/d to be optimal for outpatient programmes and only 31% thought optimal weight gain is slower in outpatient programmes than inpatient, despite this being the norm [21, preprint]. Two participants advocated for reduced dose of therapeutic food, especially towards the end of treatment. This was based on criticism of WHO’s estimation of energy requirements [22, 23] and non-inferiority as reflected in the OptiMA-DRC trial [24].

Differing perceptions reflects insufficient evidence on this topic. Whilst proving causality between PMWG and longer-term NCD is difficult because long-term RCTs are complex and costly, novel methods and approaches using biomarkers might help inform future policy/practice [25]. Furthermore, considering that programme-level weight gain in therapeutic feeding programmes is slower now than historically, too rapid PMWG may pose less of a potential threat [21, preprint].

Almost all participants (97%) believed that improving child development is an important/very important medium-term aim of treatment programmes, with 22% even ranking this as the most important aim. With increasing availability of assessment tools, future research (and potentially programmes) should consider measuring child development [26–28].

### Strengths and limitations

Our strength is that we included participants from diverse settings and professional backgrounds, ensuring our findings are broadly applicable across various contexts. This diversity strengthens the relevance and generalisability of our results. However, we also acknowledge limitations. Some respondents were excluded as they had not adequately provided consent. These individuals may have been similar (*e.g.,* time-constrained professionals), meaning a key demographic may have been excluded. Small sample size may affect generalisability and robustness of results. Another limitation is that study consent/information sheets introduced participants to our project aims, which may have influenced their answers. Confirmation bias may have impacted results. However, the substantial variability of our findings indicates that confirmation bias was moderate. There may also have been selection bias in interview participation, in that people with strong views are more likely to make themselves available. While the wealth of data we collected is undoubtedly a strength, it was impossible to present nuances of all opinions. However, thematic analysis picked up the most frequently occurring opinions.

## CONCLUSIONS

There is a large variation in the perceptions of key stakeholders on optimal PMWG, and the potential for treatment programmes to impact long-term NCD risk. While most stakeholders believe that we can optimise treatment programmes to maximise long-term outcomes, some are concerned that short-term outcomes may suffer. Better future data is urgently needed so that practice is based on evidence rather than opinion.

## Data Availability

Anonymised data will be made available on the project data repository: https://datacompass.lshtm.ac.uk/id/eprint/2655/.

## ACKNOWLEDGEMENTS

Thanks to all the survey and interview participants.

## CONTRIBUTORS

MK had the initial concept for this study and was responsible for academic supervision and project administration. All authors contributed to details of ethics and planning. GOD launched the online survey, organised and conducted interviews, and conducted the analysis. All authors were involved in survey distribution. GOD drafted the first version of the manuscript. All authors reviewed and commented on the interpretation of the findings.

