## Supplementary Materials for "Perceptions of Optimal Post-Malnutrition Weight Gain & Growth: a global mixed methods study"

### Perceptions and understandings of Post-Malnutrition Weight Gain & Growth: a qualitative survey

#### Survey Information Sheet

Please read this information sheet carefully before proceeding to the consent form on the next page.

**Introduction**

You are invited to take part in a mixed-methods research study investigating current perceptions of post-malnutrition weight gain and growth. The following sections provide information on the purpose of the study and what participation in the study will involve. Please read this page carefully before providing your informed consent.

If you require any further information, please do not hesitate to contact the research team (at). You may speak to others about the study if you wish.

**What is the purpose of the study?**

This study contributes to a work package within the Medical Research Council's "CHANGE" project (Child malnutrition & Adult NCDs: Generating new Evidence on mechanistic links in Jamaica, Malawi & Ethiopia to inform future policy & practice). The overall aim of the CHANGE project is to "optimise severe malnutrition treatment programmes by better understanding the mechanisms linking infant/child undernutrition to longer-term (adult) NCD".

This present study aims to understand current perceptions of post-malnutrition weight gain and growth, and its relation to risks of non-communicable diseases (NCDs). As a descriptive mixed-methods study, our findings will be used to generate hypotheses and inform future intervention studies on this topic.

**Why have I been asked to take part?**

We would like to gather perceptions of individuals working in the field of severe malnutrition or child health. You have been invited to take part becaue we are interested in recruiting individuals from a range of backgrounds and relevant sectors.

**Do I have to take part?**

No; Your participation is entirely voluntary. If you wish to take part in the study, we will ask you to provide your electronic consent by ticking a consent box on the next page, before commencing the survey.

**Can I change my mind about taking part?**

Yes; You may refuse to participate or withdraw from the study at any time without having to provide any reason.

**What will I have to do?**

You will complete an online survey which is expected to take approximately 10-15 minutes. At the start of the survey, you will be asked brief questions about your professional background. The main survey involves short answer questions and multiple-choice questions regarding your perceptions on (1) optimal rates of growth post-malnutrition and (2) aims of malnutrition treatment programmes.

At the end of the online survey, you will have the option to participate in an in-depth interview to help us further understand your perceptions of post-malnutrition growth, aims of malnutrition treatment programmes, and their relations to long-term risks of non-communicable diseases. If you are willing to be contacted for this interview, you will be asked to provide your email address and your electronic consent at the end of the survey. Only a small number of participants will be approached to participate in the interview. The interview will take place online, via Zoom audio or video call, and is expected to last approximately 30-45 minutes.

**What are the possible risks and disadvantages?**

As we will not be collecting any sensitive or personal information, we do not foresee any risks or disadvantages in taking part in the study. We would like to highlight that these questions are designed to gather your opinions on the topic. There are no right or wrong answers; **All responses are valid and important to us**. Your answers will only represent your individual views and not that of any organisation you are associated with.

**What are the possible benefits?**

Findings obtained from the study will help provide a better understanding of current perceptions of post-malnutrition weight gain and growth, and may inform hypotheses and designs of future studies on this topic. This study may not provide any direct benefits for you personally.

**What if something goes wrong?**

If you have any concerns about this study, you may speak to the Lead Investigator, Dr. Marko Kerac, whose research team will do their best to answer your queries: (or). If you remain unhappy and wish to file a formal complaint, you may contact Patricia Henley at or +44 (0) 20 7927 2626.

The London School of Hygiene and Tropical Medicine (LSHTM) holds insurance policies which apply to this study. If you experience harm or injury as a result of taking part in this study, you may be eligible to claim compensation.

**What will happen to information collected about me?**

All data and information collected from you will be securely kept in protected, encrypted files. Only the staff involved in the study and regulatory authorities performing monitoring, audits, or inspections for the study will be allowed to access the data and information collected.

The only piece of personal information we will request from you will be your email address, if you are willing to take part in the online interview. All email correspondence will be deleted from our records once we have received all consent forms and have scheduled the interviews. Audio/video recordings of interviews will be deleted once the content has been transcribed. You may request to review your interview transcript if desired.

All survey data and interview transcripts will be anonymised and de-identified.

At the end of the project, the study data will be archived at LSHTM's digital research repository (LSHTM Data Compass https://datacompass.lshtm.ac.uk/information.html). Your personal information will not be included and the data will be anonymous and unidentifiable.

**Where can you find out more about how your information is used?**

You can find out more about how we use your information:

- at https://www.lshtm.ac.uk/sites/default/files/research-participant-privacy-notice.pdf
- by asking one of the research team via (or)

**What will happen to the results of this study?**

Results of the study may be published in a relevant journal. Your personal information will not be included in the study report. All data will be anonymised and de-identified upon publication.

**Who is organising this study?**

LSHTM is organising the study. As the Data Controller for the study, LSHTM is responsible for the proper collection, storage, and analysis of your data.

**Who has reviewed this study?**

This study has been reviewed and approved by the LSHTM Research Ethics Committee to protect your interests.

**Further information and contact details**

Thank you for taking time to read this information sheet. If you would like to take part in this study, please proceed to the next page to provide your electronic consent. The online survey will commence after you have provided your electronic consent. If you would like any further information, please contact the study's research team at.

#### Survey Consent form

If you would like to take part in the following study, "Perceptions and understandings of Post-Malnutrition Weight Gain & Growth: a survey and key informant interview study", please provide your electronic consent by ticking the following statements. *

- [ ] I confirm that I have read and understood the information sheet for this study. I have had the opportunity to consider the information and ask questions to appropriate personnel, as stated on the information sheet. Any questions I have asked have been answered satisfactorily.
- [ ] I understand that my consent is voluntary and that I am free to withdraw this consent at any time without giving any reason.
- [ ] I understand that data collected during the study may be looked at by authorised individuals where it is relevant to my participation in this research. I give permission for these individuals to have access to these records.
- [ ] I understand that data collected from me or about me may be shared on a public data repository and/or with other researchers, and that I will not be identifiable from this information.
- [ ] I understand that data collected during this study may be used in research publications, and that all data will be anonymised and unidentifiable.
- [ ] I agree to participating in this study.

To support our research findings with qualitative evidence, we may directly quote statements given by participants in their online survey responses or interview transcripts. These quotes will be anonymised and you will not be identifiable from these quotes.

- [ ] I give permission for any written or transcribed statements recorded from the survey or interview to be directly quoted in publication as anonymised, de-identified statements.

Please provide your electronic signature by filling in your full name in the text box below. *

#### Participant's Professional Background

What type of organisation do you work for? *

- ◯ Non-Governmental Organisation (NGO)/Charity
- ◯ Policy development
- ◯ Government
- ◯ United Nations
- ◯ Healthcare
- ◯ Public Health
- ◯ Academic
- ◯ Independent
- ◯ Other

If you selected Other, please specify: *

____________________

What is your professional role? (Select all that apply) *

- ☐ Doctor (paediatrics)
- ☐ Doctor (other)
- ☐ Nurse
- ☐ Nutritionist
- ☐ Mental Health Specialiist
- ☐ Public Health Specialist
- ☐ Programme manager
- ☐ Other

If you selected Other, please specify: *

How long have you been working in this role or in similar work? *

- ◯ 0-5 years
- ◯ 6-10 years
- ◯ 11-15 years
- ◯ 16-20 years
- ◯ 20+ years

Which geographical region is your work focused on? (Select all that apply) *

- ☐ Global
- ☐ North Africa
- ☐ Sub-Saharan Africa
- ☐ Latin-America & Carribean
- ☐ North America
- ☐ Central Asia
- ☐ East Asia
- ☐ South-East Asia
- ☐ South Asia
- ☐ West Asia
- ☐ Europe
- ☐ Oceania

What context do you work in? (Select all that apply) *

- ☐ Humanitarian
- ☐ Development
- ☐ Other

If you selected Other, please specify: *

#### Perceptions of Post-Malnutrition Weight Gain and Growth: In-patient setting

Malnutrition treatment programmes monitor a child's progress by measuring the child's rate of weight gain and growth as they receive therapeutic feeds.

The rate of daily weight gain for a child is often recorded as the **number of grams gained per kilogram of child's weight per day (g/kg/day).**

Malnutrition treatment programmes can be delivered via in-patient settings and/or community outpatient services. Follow-up growth monitoring **after** the child is discharged from the malnutrition treatment programme is also common.

For the following two questions, please provide the answer which best describes your opinion of an optimal rate of daily weight gain in that particular setting.

What would you consider an optimal rate of weight gain during the IN-PATIENT phase of malnutrition treatment? (in g/kg/day) *

Optimal rate of weight gain in g/kg/day

- ◯ 0-5
- ◯ 5-10
- ◯ 10-15
- ◯ 15-20
- ◯ 20+

#### Perceptions of Post-Malnutrition Weight Gain and Growth: Out-patient or community-based setting

What would you consider an optimal rate of weight gain during the OUT-PATIENT phase of malnutrition treatment or a CMAM programme (Community-based Management of Acute Malnutrition)? (in g/kg/day) *

Optimal rate of weight gain in g/kg/day

- ◯ 0-5
- ◯ 5-10
- ◯ 10-15
- ◯ 15-20
- ◯ 20+

#### Perceptions of Post-Malnutrition Weight Gain and Growth: Post-discharge

Please study the following two Weight-for-Age Z-score (WAZ) growth charts carefully and answer the questions below.

Blue dots on the chart represent the child's growth. The x-axis shows age.

Solid black/red/green lines on the chart are the World Health Organisation (WHO) Weight-for-Age Z-score (WAZ) thresholds. The green line is the median.

NB: The **starting WAZ (at discharge from treatment programme)** are **the same** in both charts.

*You may right click on the charts to open in a new window on a desktop/laptop. On a smartphone, please zoom in.*

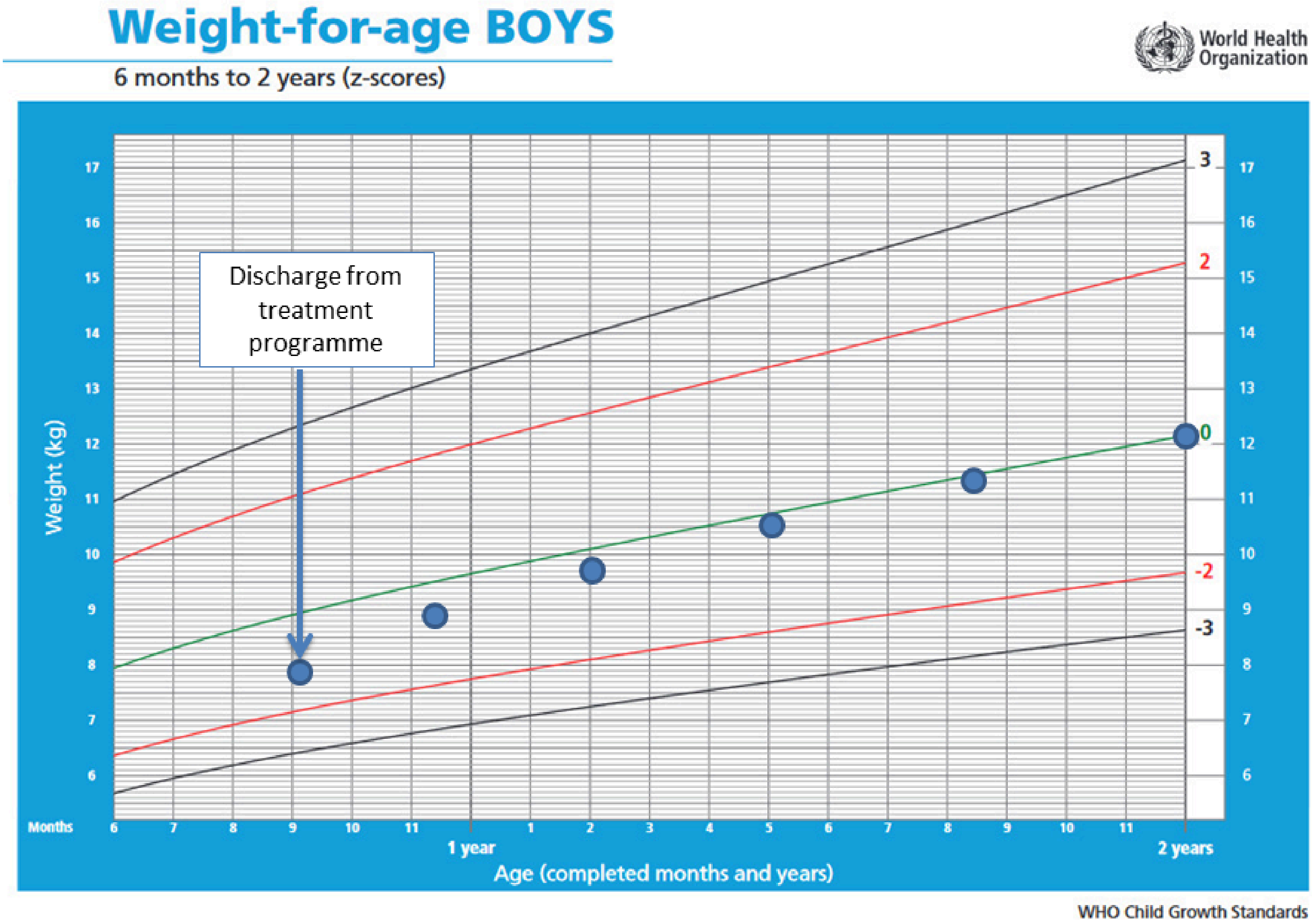


Chart A

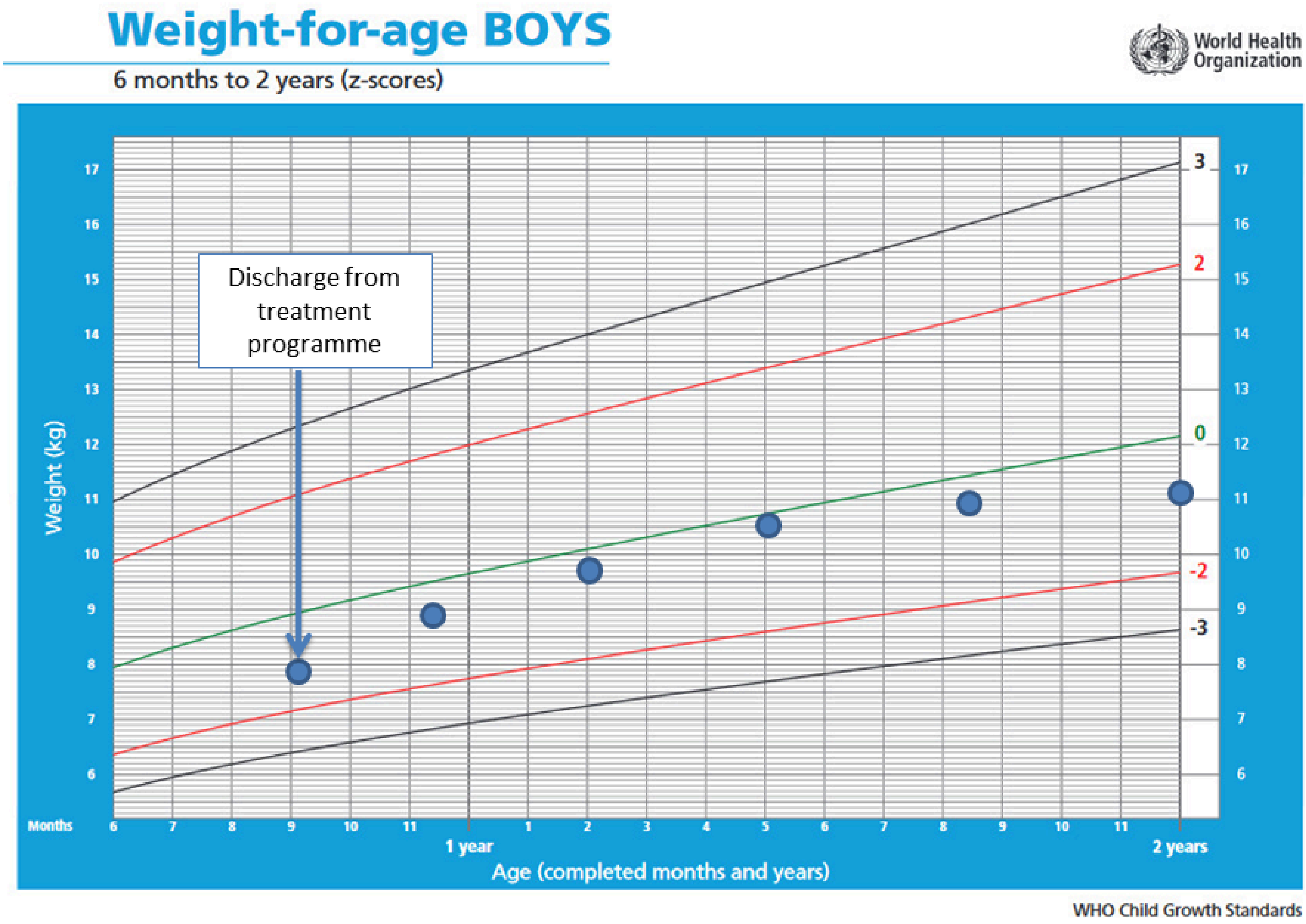


Chart B

In your opinion, which growth chart shows a more optimal (healthier/more desirable) growth pattern? Please consider the (i) starting WAZ, (ii) ending WAZ, (iii) rates of growth relative to WHO z-score thresholds, and (iv) trajectory of these growth patterns. *

- ◯ Chart A
- ◯ Chart B
- ◯ No difference

Please briefly explain WHY the growth pattern you have chosen is more optimal (healthier/more desirable)? NB: Please avoid simply describing the growth pattern.

**Even a few words on your choice would be helpful!**

#### Perceptions of Post-Malnutrition Weight Gain and Growth: Post-discharge

Please study the following two Weight-for-Age Z-score (WAZ) growth charts carefully and answer the questions below.

Blue dots on the chart represent the child's growth. The x-axis shows age.

Solid black/red/green lines on the chart are the World Health Organisation (WHO) Weight-for-Age Z-score (WAZ) thresholds. The green line is the median.

NB: The **WAZ at the start (at admission to malnutrition treatment programme) and at the end (post-discharge, aged 2 years)** are **the same** in both charts.

*You may right click on the charts to open in a new window on a desktop/laptop. On a smartphone, please zoom in.*

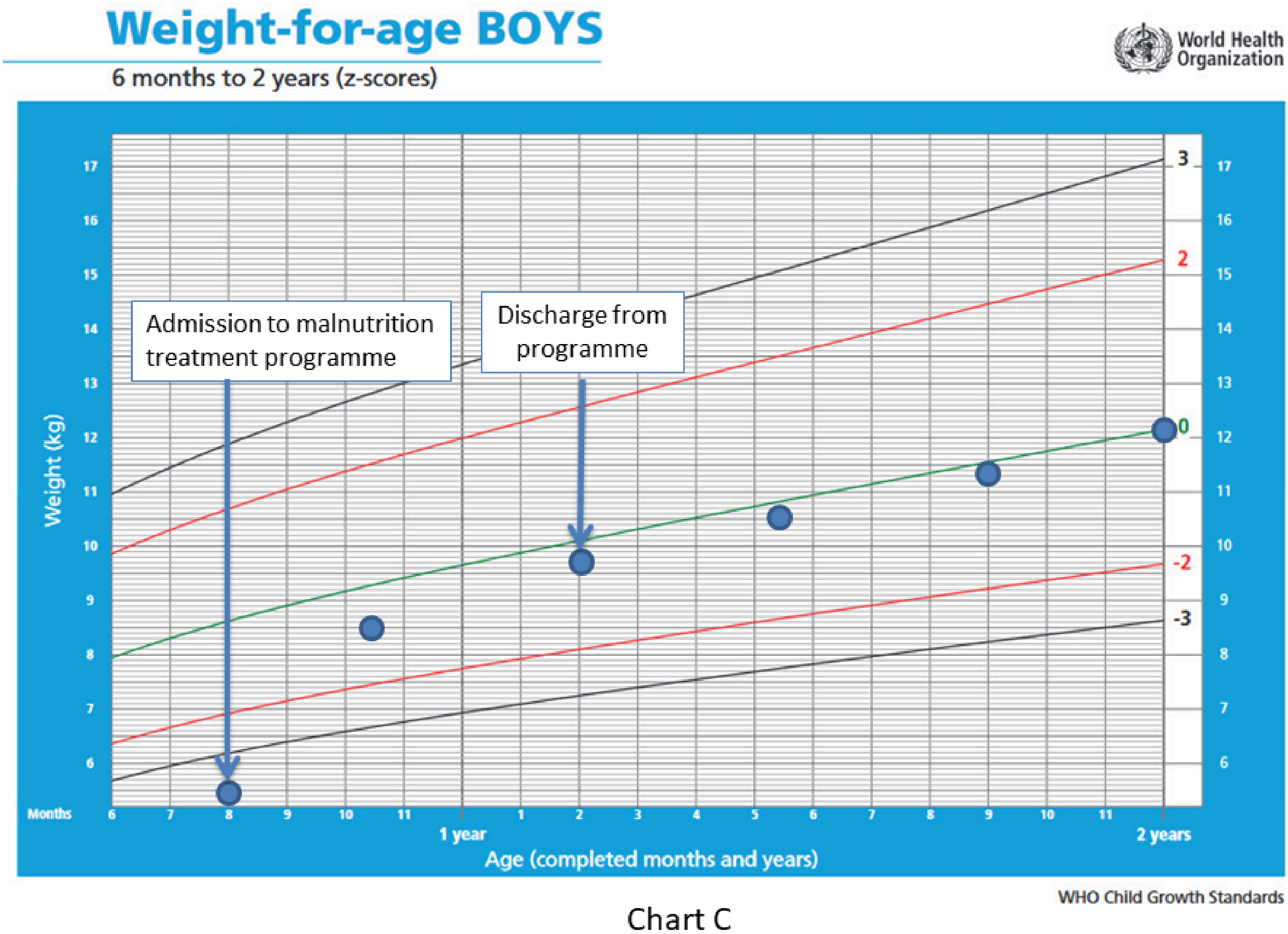


Chart C

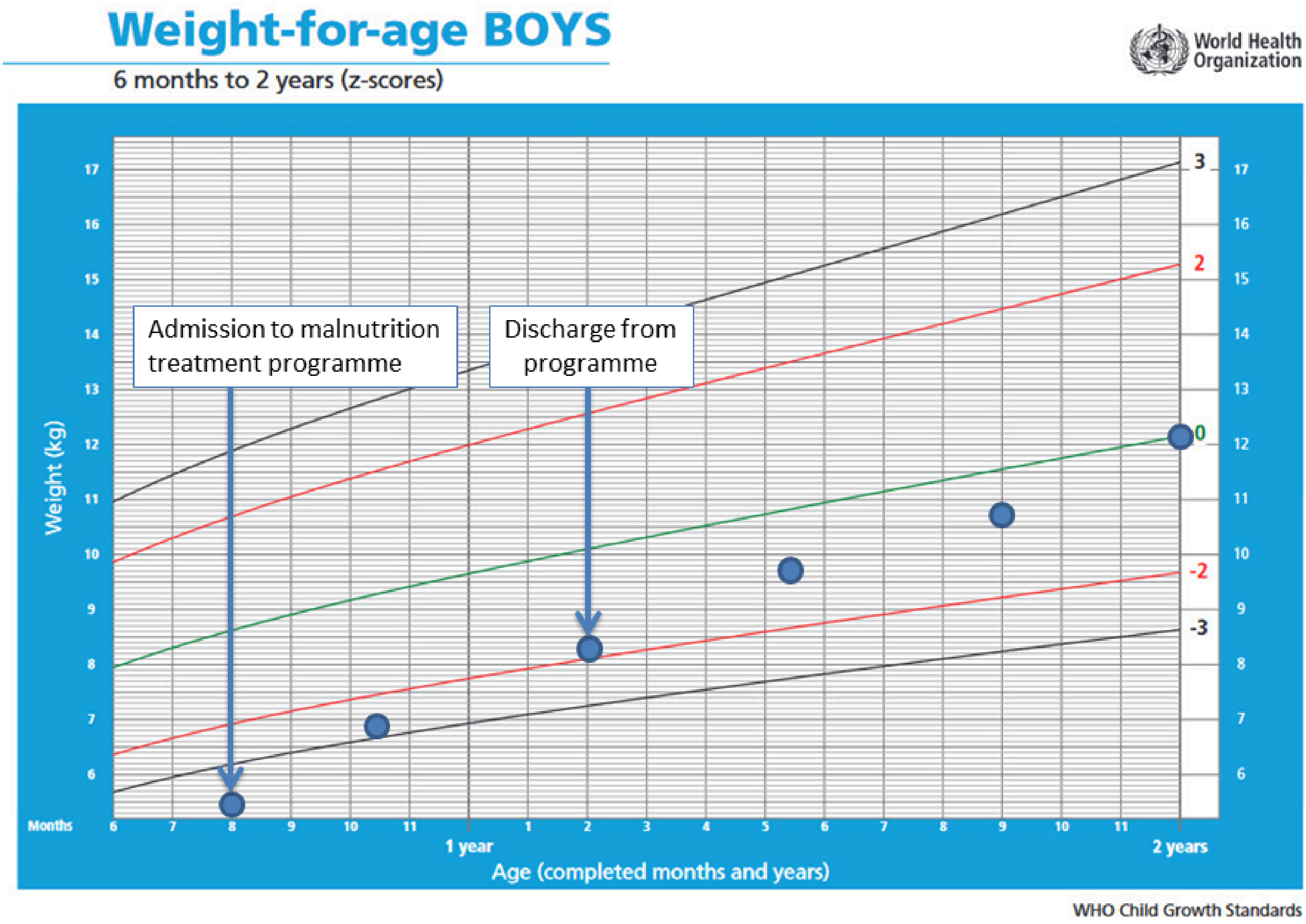


Chart D

In your opinion, which growth chart shows a more optimal (healthier/more desirable) growth pattern? Please consider the (i) starting WAZ, (ii) ending WAZ, (iii) rates of growth relative to WHO z-score thresholds, and (iv) trajectory of these growth patterns. *

- ◯ Chart C
- ◯ Chart D
- ◯ No difference

Please briefly explain WHY the growth pattern you have chosen is more optimal (healthier/more desirable)? NB: Please avoid simply describing the growth pattern.

**Even a few words on your choice would be helpful!**

#### Perceptions of Post-Malnutrition Weight Gain and Growth: Post-discharge

Please study the following two Weight-for-Age Z-score (WAZ) growth charts carefully and answer the questions below.

Blue dots on the chart represent the child's growth. The x-axis shows age.

Solid black/red/green lines on the chart are the World Health Organisation (WHO) Weight-for-Age Z-score (WAZ) thresholds. The green line is the median.

NB: The **starting WAZ (at admsision to the malnutrition treatment programme)** are **the same** in both charts.

*You may right click on the charts to open in a new window on a desktop/laptop. On a smartphone, please zoom in.*

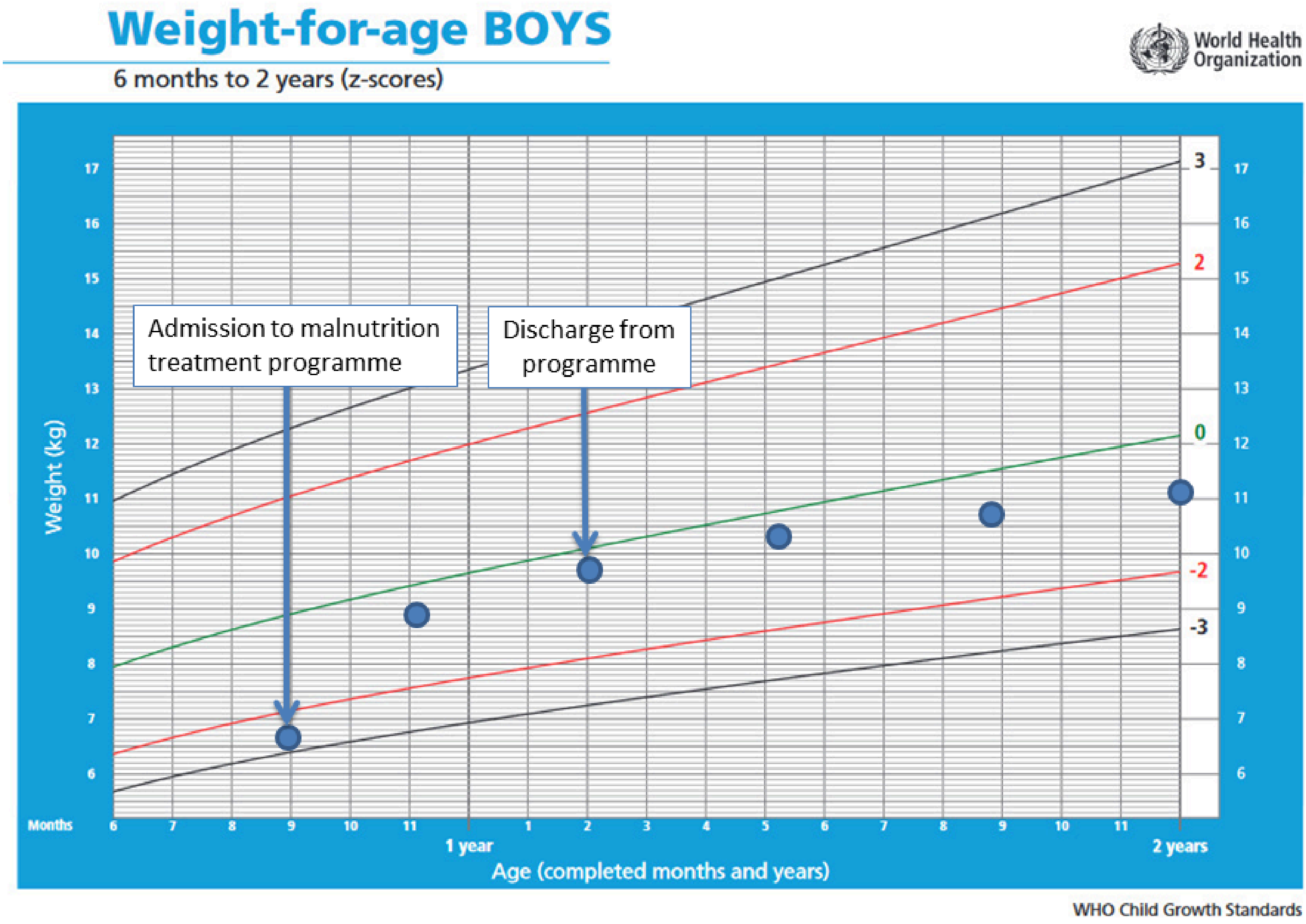


Chart E

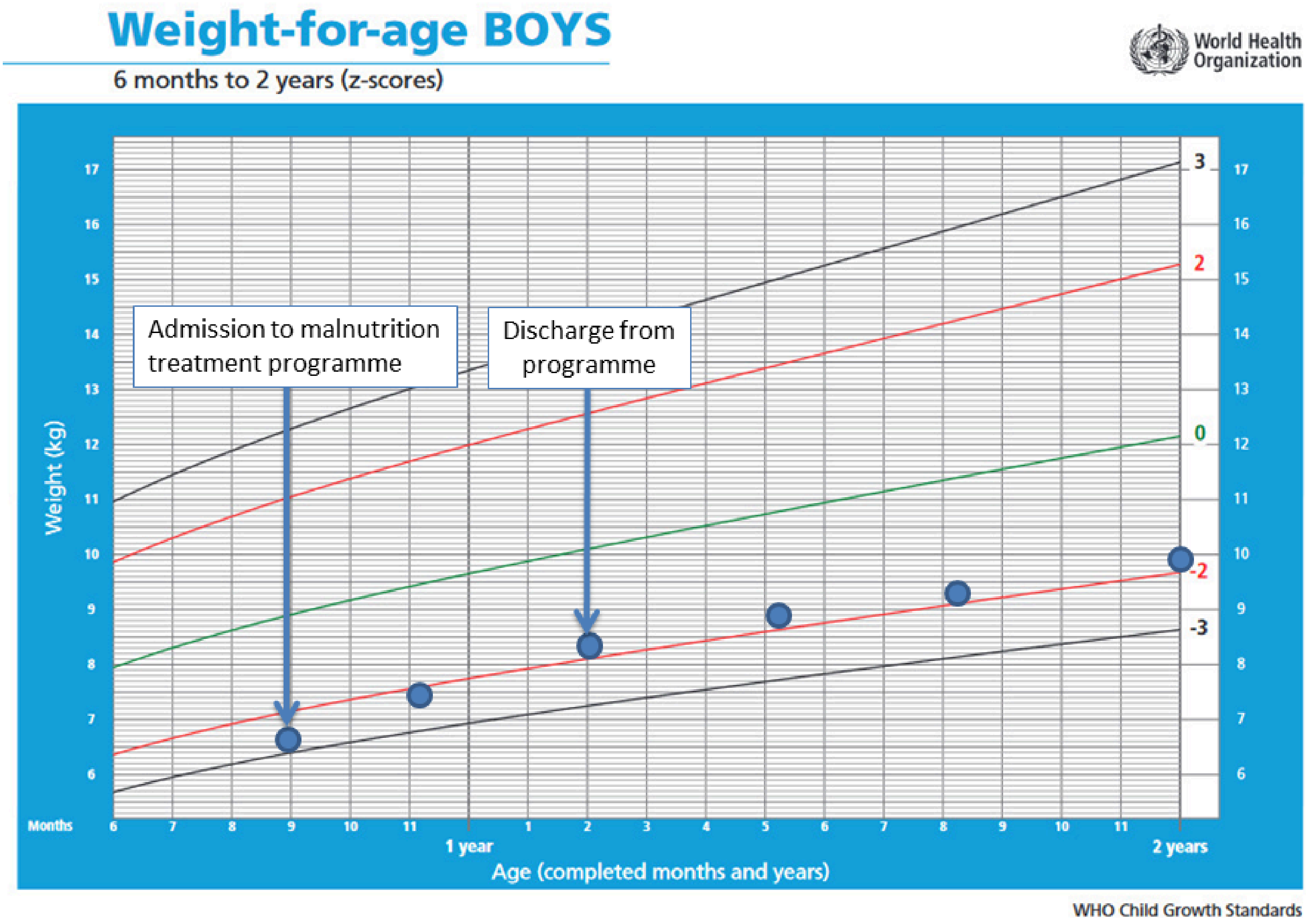


Chart F

In your opinion, which growth chart shows a more optimal (healthier/more desirable) growth pattern? Please consider the (i) starting WAZ (ii) ending WAZ, (iii) rates of growth relative to WHO z-score thresholds, and (iv) trajectory of these growth patterns. *

- ◯ Chart E
- ◯ Chart F
- ◯ No difference

Please briefly explain WHY the growth pattern you have chosen is more optimal (healthier/more desirable)? NB: Please avoid simply describing the growth pattern.

**Even a few words on your choice would be helpful!**

#### Perceptions of Post-Malnutrition Weight Gain and Growth: Post-discharge

Please study the following two **HEIGHT**-for-Age Z-score (HAZ) growth charts carefully and answer the questions below.

Blue dots on the chart represent the child's growth. The x-axis shows age.

Solid black/red/green lines on the chart are the World Health Organisation (WHO) Height-for-Age Z-score (HAZ) thresholds. The green line is the median.

NB: The **ending HAZ** are **the same** in both charts.

*You may right click on the charts to open in a new window on a desktop/laptop. On a smartphone, please zoom in.*

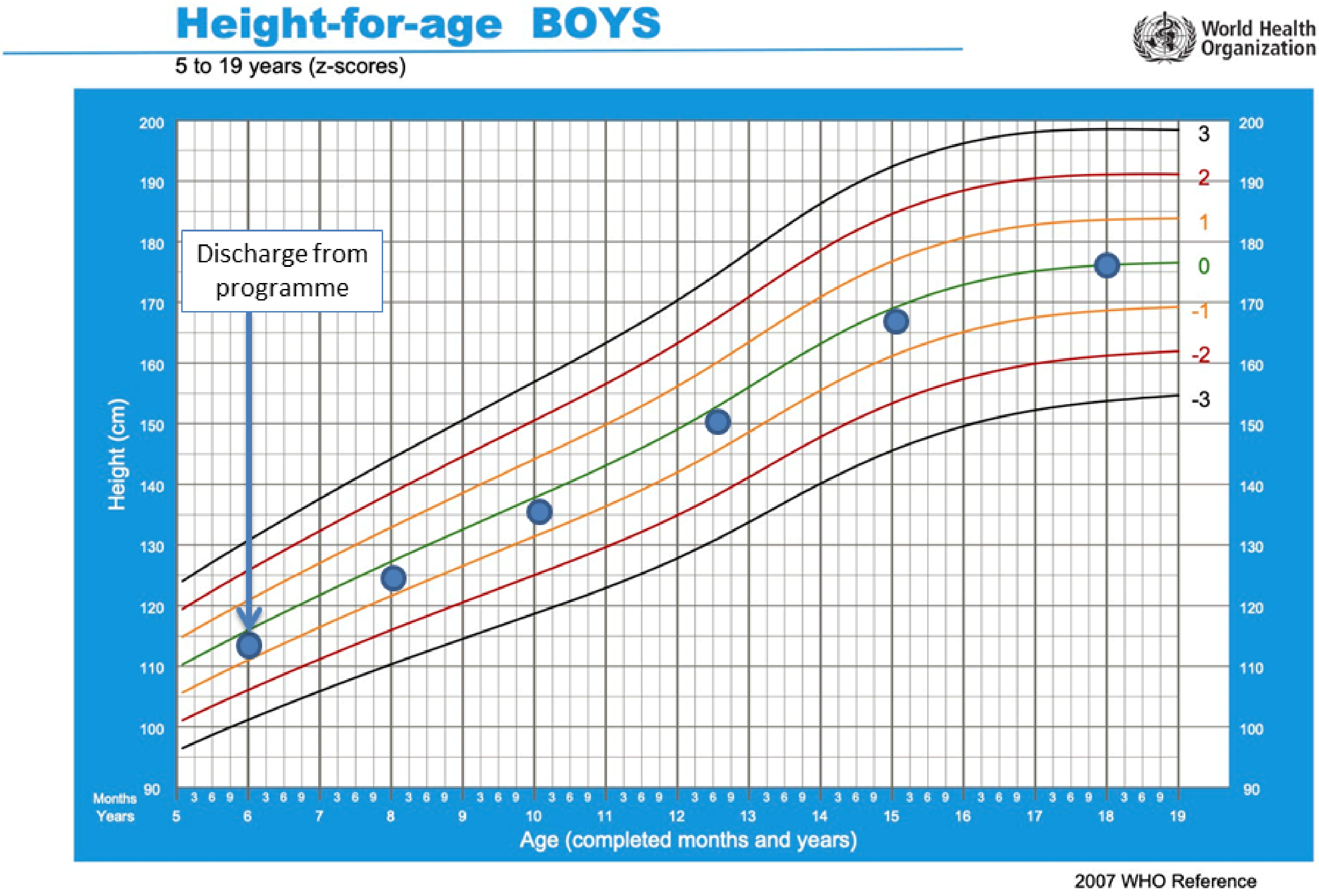


Chart G
(HAZ)

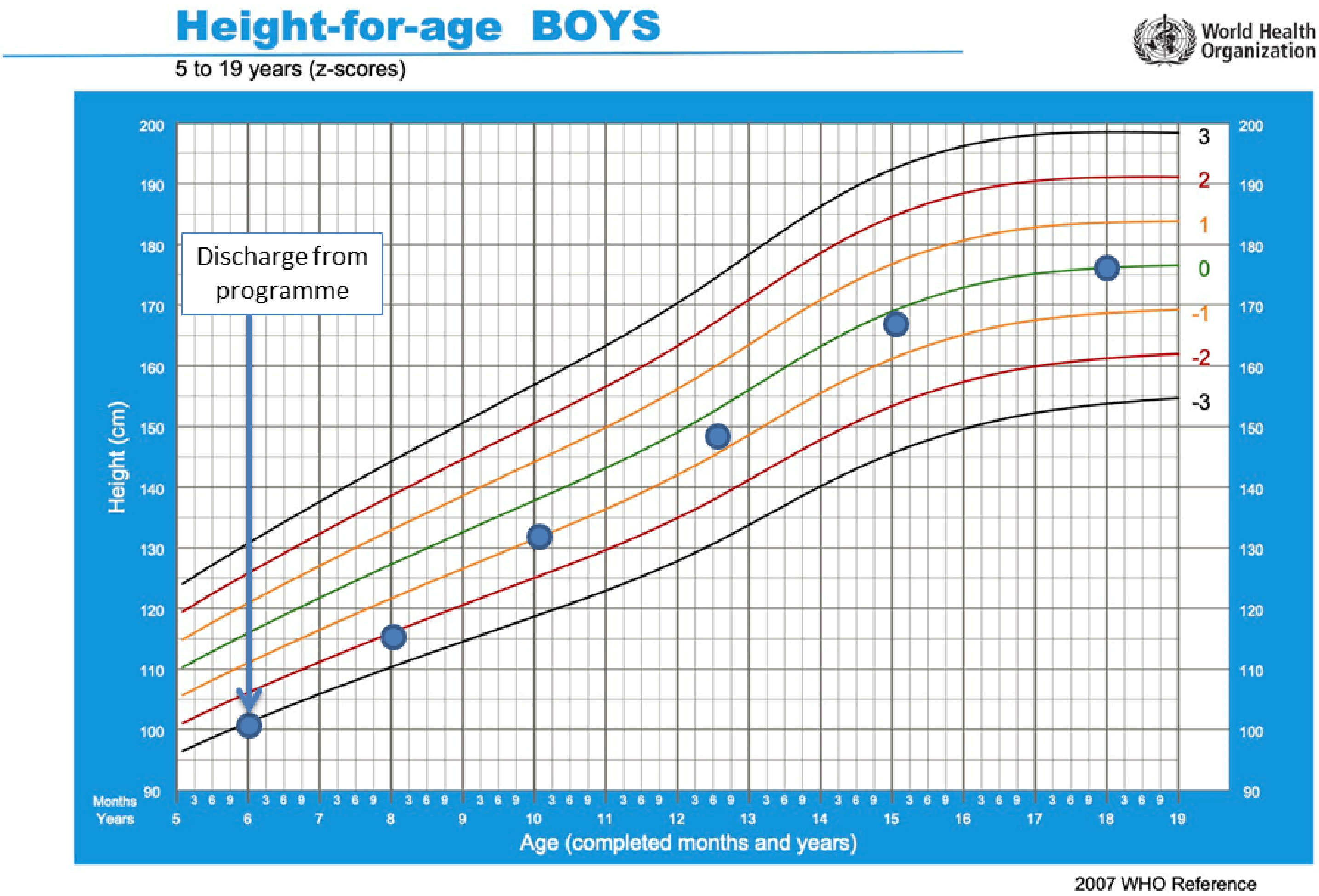


Chart H
(HAZ)

In your opinion, which growth chart shows a more optimal (healthier/more desirable) growth pattern? Please consider the (i) starting HAZ, (ii) ending HAZ, (iii) rates of growth relative to WHO z-score thresholds, and (iv) trajectory of these growth patterns. *

- ◯ Chart G (HAZ)
- ◯ Chart H (HAZ)
- ◯ No difference

Please briefly explain WHY the growth pattern you have chosen is more optimal (healthier/more desirable)? NB: Please avoid simply describing the growth pattern.

**Even a few words on your choice would be helpful!**

#### Perceptions of Post-Malnutrition Weight Gain and Growth: Post-discharge

Please study the following two **HEIGHT**-for-Age Z-score (HAZ) growth charts carefully and answer the questions below.

Blue dots on the chart represent the child's growth. The x-axis shows age.

Solid black/red/green lines on the chart are the World Health Organisation (WHO) Height-for-Age Z-score (HAZ) thresholds. The green line is the median.

NB: The **starting HAZ (at discharge from malnutrition treatment programme)** and **ending HAZ** are **DIFFERENT** in both charts.

*You may right click on the charts to open in a new window on a desktop/laptop. On a smartphone, please zoom in.*

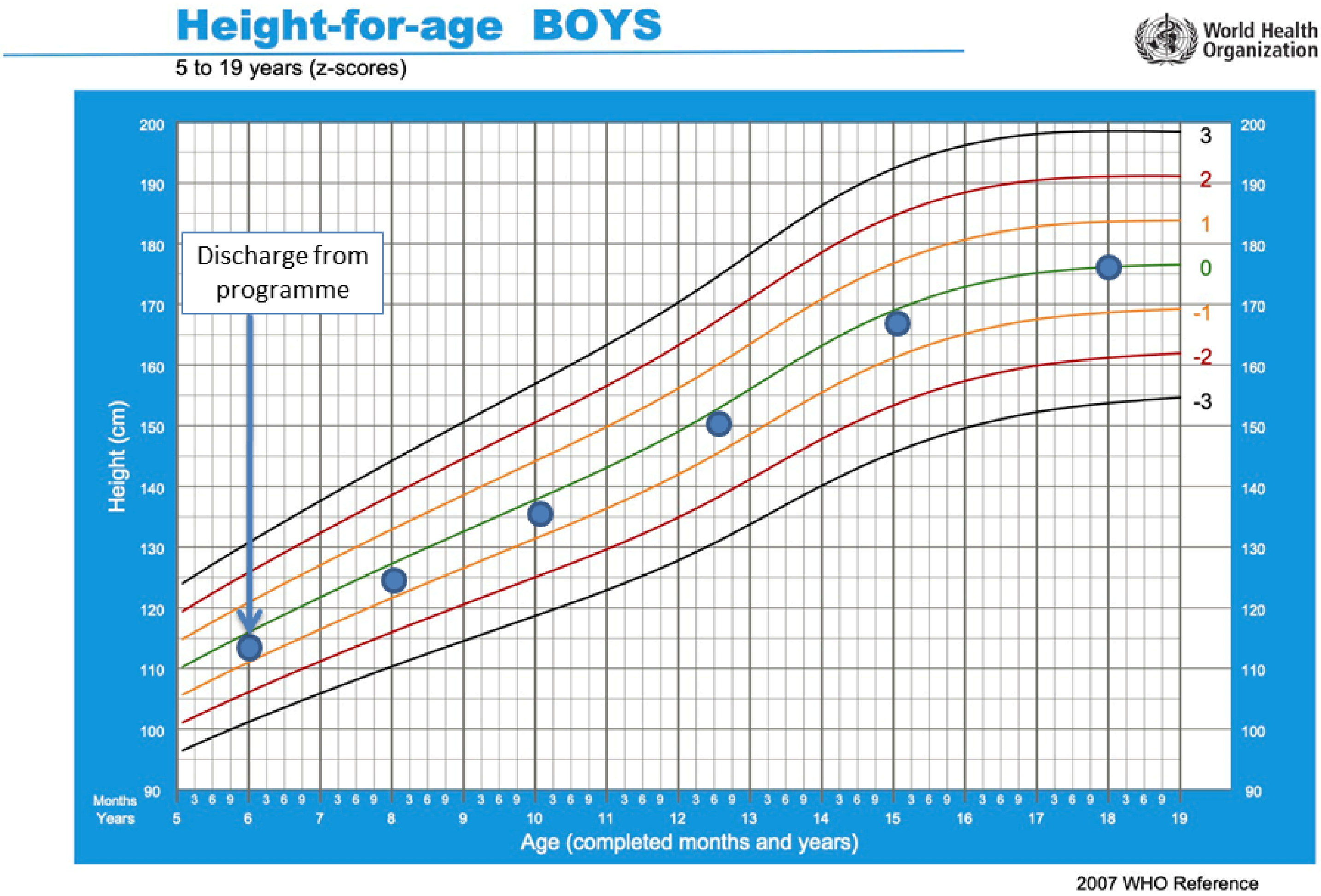


Chart J
(HAZ)

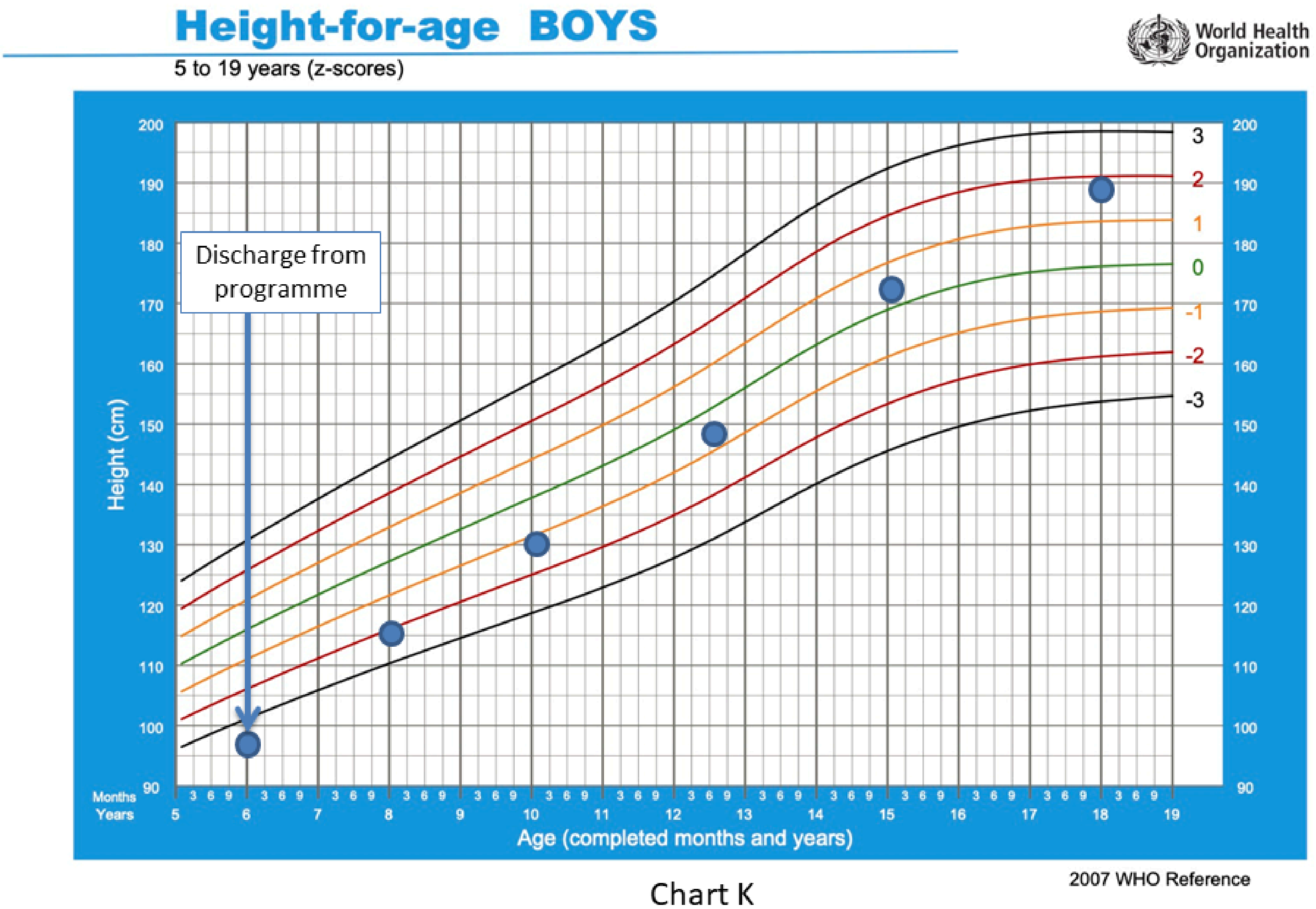


Chart K
(HAZ)

In your opinion, which growth chart shows a more optimal (healthier/more desirable) growth pattern? Please consider the (i) starting HAZ, (ii) ending HAZ, (iii) rates of growth relative to WHO z-score thresholds, and (iv) trajectory of these growth patterns. *

- ◯ Chart J (HAZ)
- ◯ Chart K (HAZ)
- ◯ No difference

Please briefly explain WHY the growth pattern you have chosen is more optimal (healthier/more desirable)? NB: Please avoid simply describing the growth pattern.

**Even a few words on your choice would be helpful!**

#### Short- and long-term aims of malnutrition treatment programmes

There is growing evidence that childhood malnutrition influences health outcomes and development in the short- and long-term.

##### In your opinion, how important is "Improving child development" as a medium-term aim of child malnutrition treatment programmes? *

Improving child development

- ◯ Very important
- ◯ Important
- ◯ Moderately important
- ◯ Of low importance
- ◯ Not at all important

##### In your opinion, how important is "Reducing the risk of non-communicable diseases in adulthood" as a long-term aim of child malnutrition treatment programmes? *

Reducing the risk of non-communicable diseases in adulthood

- ◯ Very important
- ◯ Important
- ◯ Moderately important
- ◯ Of low importance
- ◯ Not at all important

#### Short- and long-term aims of malnutrition treatment programmes

Do you believe that malnutrition treatment programmes have a role in reducing the risk of adulthood non-communicable diseases, given that children survive childhood malnutrition? *

- ◯ Yes
- ◯ No
- ◯ Don't know

Please briefly explain your answer.

**Even a few words on your choice would be helpful!**

#### Short- and long-term aims of malnutrition treatment programmes

The following five outcomes are all potential short- and long-term aims of malnutrition treatment programmes. Please **rank** the following five aims in order of importance and priority in the context of malnutrition treatment programmes. (1 = most important, 5 = least important).

**Please do not give the same ranking to more than one aim.**

NB: There are no right or wrong answers. We simply want to gather your personal opinions and preferences. Aims are listed in alphabetical order.

*

Improving child development*

- ☐ 1 (most important)
- ☐ 2
- ☐ 3
- ☐ 4
- ☐ 5 (least important)

Preventing mortality

- ☐ 1 (most important)
- ☐ 2
- ☐ 3
- ☐ 4
- ☐ 5 (least important)

Reducing the risk of non-communicable diseases in adulthood

- ☐ 1 (most important)
- ☐ 2
- ☐ 3
- ☐ 4
- ☐ 5 (least important)

Reducing the risk of short-term morbidity**

- ☐ 1 (most important)
- ☐ 2
- ☐ 3
- ☐ 4
- ☐ 5 (least important)

Reducing the risk of stunting by 2 years of age

- ☐ 1 (most important)
- ☐ 2
- ☐ 3
- ☐ 4

☐ 5 (least important)

*Improving child development may include improving educational potential and preventing disability

**Short-term morbidity, e.g. diarrhoea, pneumonia, acute infections, or illnesses

### Understanding your perceptions of Post-Malnutrition Weight Gain and Growth

Do you think that slower rate of daily weight gain and growth post-malnutrition (e.g. by providing a lower-energy therapeutic feed*) could be beneficial for the child in any way?

* e.g. which could be achieved by prescribing feeds at 150 kcal/kg/day (i.e. lower end of the WHO-recommended range), rather than the 200 kcal/kg/day (i.e. top end of range)

*

- ◯ Yes
- ◯ No
- ◯ Don't know

Please briefly explain your answer.

**Even a few words on your choice would be helpful!**

Any further comments regarding topics covered in this survey?

#### End of Online Survey

Thank you for completing the online survey.

If you are willing to take part in the next stage of the study (the in-depth online interview lasting approximately 30-45 minutes via Zoom), please leave your email address below. Otherwise, leave the space blank.

By providing your full name in the space below, you give permission to be contacted via email to schedule the online interview.
